# The effects of a dietary supplement on brain function and structure in Junior A ice hockey players: a prospective randomized trial

**DOI:** 10.1101/2023.04.03.23288079

**Authors:** Logan T. Breuer, Shaun D. Fickling, David A. Krause, Ryan C.N. D’Arcy, Tory O. Frizzell, Tania Gendron, Michael J. Stuart

**Affiliations:** Department of Orthopedic Surgery, Division of Sports Medicine, Mayo Clinic, Rochester, MN, 55905, USA; Centre for Neurology Studies, HealthTech Connex, Metro Vancouver, BC, V3V0C6, Canada; Department of Physical Medicine and Rehabilitation, Program in Physical Therapy, Mayo Clinic, Rochester, MN, 55905, USA; BrainNET, Health and Technology District, Surrey, BC, V3V0C6, Canada; DM Centre for Brain Health, Radiology, University of British Columbia, Metro Vancouver, BC, V6T1Z4, Canada; Department of Neuroscience, Mayo Clinic, Jacksonville, FL 32224, USA; Mayo Clinic Graduate School of Biomedical Sciences, Mayo Clinic, Jacksonville, FL 32224, USA

## Abstract

**Background:** Neurocognitive impairment linked to head impact exposure in otherwise healthy, non-concussed athletes may be associated with adverse long-term outcomes. The primary purpose of this study was to evaluate whether a dietary supplement, Synaquell^TM^, supports brain function and structure in male Junior A ice hockey players over the course of a season.

**Methods:** Players underwent pre-season testing, were randomized into a placebo or dietary supplement group, then were retested after the season. Objective tests included: NeuroCatch^®^ portable evoked potential platform, King-Devick Test of rapid number naming, and blood biomarker assay for neurofilament light chain (NfL).

**Results:** Multivariate analysis revealed significant differences in neurocognitive changes between groups from pre to postseason after controlling for covariates related to head impact exposure. Post-hoc tests showed significant within-subject differences between groups from pre- to post-season in both N100 latency (p = 0.005) and King-Devick score (p = 0.043). Univariate tests of the NeuroCatch results replicated prior findings of a N400 amplitude decrease (p = 0.017) and N100 latency increase (p = 0.049) in the placebo group, but not in the dietary supplement group.

**Conclusions:** This prospective, randomized trial showed that, compared to the placebo group, a multi-ingredient dietary supplement significantly affected objective measures of brain function and structure in Junior A ice hockey players from pre- to post-season. Further investigation into the effects of dietary supplementation on the contact athlete’s brain is warranted.

## Introduction

Athletes sustaining repetitive body and head impacts may experience impaired neurocognitive function without observable symptoms (Bailes et al. 2013; Mainwaring et al. 2018). These impacts with resultant force transmission to the brain occur frequently over the course of a season in contact sports, such as ice hockey. Objective neurocognitive tests have shown pre- and post-season differences in athletes exposed to repetitive brain forces in the absence of a clinically diagnosed concussion (Abbas et al. 2015; Talavage et al. 2014; Tsushima et al. 2016). The emergence of detectable neurocognitive impairments is of critical concern due to recent evidence linking impact exposure to progressive neurodegenerative disease. The risk of chronic traumatic encephalopathy (CTE) has been shown to double with every 2.6 years of contact exposure (Mez et al., 2020).

Proper medical evaluation is imperative to assessing the effects of repetitive head impacts (Pender et al. 2020). Advances have been made in point-of-care evaluation with objective neurophysiological measures, oculomotor tests, and neuro-axonal biomarkers. The NeuroCatch^®^ is a medical device used to evaluate objective neurocognitive changes within a brain vital signs framework (Ghosh Hajra et al. 2016). Electroencephalography (EEG) data are processed to quantify event-related potential (ERP) responses for the N100 (auditory sensation; Davis 1939), the P300 (basic attention; Sutton et al. 1967), and the N400 (cognitive processing; Kutas et al. 1980). The ability of the NeuroCatch^®^ to detect subtle but significant changes in cognitive brain function has led to investigations on cumulative head impact exposure and “subconcussive” impairments (Fickling et al. 2021a; Fickling et al. 2021b). Fickling et al. (2019) first detected neurocognitive impairments tracked by the N400 ERP response for cognitive processing in ice hockey athletes without a diagnosed concussion throughout the season. In a subsequent ice hockey study (Fickling et al., 2021a), neurocognitive impairments in the N400 were replicated, with additional impairments detected in the N100 ERP response for auditory sensation. The pattern of brain vital sign ERP changes over the course of the hockey season was highly correlated and linearly predictive of the number of head Impacts measured by accelerometers. These findings were replicated in youth football showing a strong linear relationship with the number of games and practices representing a general measure of impact exposure (Fickling et al., 2021b). These studies demonstrate cognitive impairments related to impact exposure occur in different sports and age groups.

Additional objective cognitive assessment tools include the King-Devick Test (K-D) and blood biomarkers. The K-D is a test of rapid number naming that relies upon saccadic function, attention, and language function. The K-D evaluates changes due to head impacts in both acute and long-term contact sports participation (Nowak et al. 2020; Munce et al. 2014; Joseph et al. 2018; Krause et al. 2021) and was shown to be reliable (Eddy et al. 2020). Caccese et al. (Caccese et al. 2019) reported a significant relationship between repetitive head impacts and change in K-D score over the course of a college football season, where greater head impact exposure was associated with poorer K-D performance. Blood biomarkers are used as an objective assay to quantify neuro-axonal damage. Neurofilament light (NfL) is a biomarker that increases in individuals exposed to repetitive head impacts (Papa et al. 2022; Oliver et al. 2016b; Oliver et al. 2018b; Wirsching et al. 2019). Rubin et al. (Rubin et al. 2019) found an association between increased serum NfL levels and increased frequencies of head impact exposure. Collectively, these three objective neurological assessments (NeuroCatch^®^, K-D, and NfL) can monitor neurocognitive changes and benchmark the efficacy of potential interventions (Smith et al. 2017; Pender et al. 2020).

A range of emerging interventions, including dietary supplements, may support brain function and structure (Walrand et al. 2021; Lucke-Wold et al. 2018; Oliver et al. 2018). Dietary supplements are non-pharmacological, easily accessible, and may support brain health (Mishra et al. 2022). A variety of supplements have been investigated in both animal models and human studies to determine their effects on the brain (Lucke-Wold et al. 2016), including magnesium (McIntosh et al. 1989; Standiford et al. 2020), resveratrol (Lopez et al. 2005, Lin et al. 2014), nicotinamide riboside (Cheng et al. 2022), curcumin (Wu et al. 2011), glutathione (Aoyama 2021), docosahexaenoic acid (DHA) (Raikes et al. 2022, Oliver et al. 2016a; Heileson et al. 2021), ketone bodies such as beta hydroxybutyrate (Lee et al. 2019), ubiquinone (Pierce et al. 2017; Pierce et al. 2018) and branched-chain amino acids such as valine, leucine and isoleucine (Aquilani et al. 2005). A combination of these ingredients and others are in the product Synaquell^TM^ (manufactured by Thorne). This dietary supplement may influence the neurometabolic cascade of concussion (Giza and Hovda, 2014) by providing critical nutrients and energy substrates to rebalance disrupted homeostatic processes (Walrand et al. 2021).

The purpose of this blinded, randomized clinical trial was to investigate if daily oral ingestion of Synaquell^TM^ supports brain function and structure over the duration of an ice hockey season. The primary hypothesis was that objective measures of brain function and structure (brain vital signs, King-Devick, NfL) would show significantly less decline as compared to a placebo control group when controlling for self-reported concussion history and number of games played (as a proxy for head-impact exposure). The secondary hypothesis was that the placebo control group would show significant impairments in brain vital sign changes from pre- to post-season, replicating analyses from prior studies evaluating contact sport athletes.

## Methods

### Participants

The study was reviewed and approved by the Institutional Review Board. Fifty-four male athletes were recruited from the North American Tier 2 Hockey League (NAHL) and the North American Tier 3 Hockey League (NA3HL). Participants were between 18 and 20 years of age (18.73 ± 0.69) at the time of enrollment, spoke fluent English, and were medically cleared to play ice hockey by the team’s medical staff. All subjects provided written consent. Exclusion criteria included allergy to the Synaquell^TM^ ingredients (Supplemental Table 1: SynaquellTM Ingredients) and contradictions for the NeuroCatch^®^ Platform (clinically documented hearing issues, hearing device, aid or implant, or an unhealthy scalp). Participants were asked to refrain from taking other dietary supplements and were excluded if they declared noncompliance. All players underwent a preseason medical evaluation and completed the Sideline Concussion Assessment Tool (SCAT5). Team rosters change during the season; therefore, participants who left their hockey team, lost medical clearance to play or did not comply with study protocols were withdrawn from the study. Athletes added to the roster during the hockey season were enrolled and tested (n=5). All participants were randomized into either the control (CON, n=28) or Synaquell^TM^ (SQ, n=26) group.

### Experimental design

This prospective, randomized clinical trial utilized repeated measures and a placebo group to determine the effects of a combination dietary supplement, Synaquell^TM^, on brain function and structure. Pre-season testing included: brain vital signs recorded using the NeuroCatch^®^ portable evoked potential platform, the K-D rapid number naming test, and venous blood analysis for NfL. Administered clinical neuropsychological questionnaires included: SCAT5, Neuro-Quality of Life Short Form Version 2.0 – Cognitive Function (nQoL), and Patient Health Questionnaire – 4 (PHQ-4). These questionnaires were repeated after the hockey season, along with the Patient Global Impression of Change (PGIC). Post-season data collection was completed with 7 days of the last regular season game.

### Brain vital signs

NeuroCatch^®^ (Version 1.1) incorporates a portable 8-channel g. Nautilus EEG system (Gtec Medical Engineering, Austria). Testing involves a 6-min automated stimulus sequence consisting of auditory tones (oddball stimuli with standard and deviant tones) interspersed with spoken prime-target word pairs (equally semantically congruent or incongruent). Participants were instructed to listen attentively, passively and remain still with their eyes fixed on a cross positioned at eye level 2 m away. Scans were conducted in a quiet, closed room away from distractions. Pre- and post-season scans were conducted in the same facility to maintain a consistent environment.

The three scalp electrodes were referenced to an electrode clipped to the right earlobe. Disposable, Ag/AgCl, adhesive electrodes were used for electro-oculogram recording from the supra-orbital ridge and outer canthus of the left eye. g. GAMMAsys electrode gel was applied to each location to ensure conductivity. Skin-electrode impedances were maintained below the standard 30kΩ threshold at each site.

Scan quality results were evaluated immediately, with all EEG data subsequently processed off-line in Python. Pre-processing was performed using a fourth-order Butterworth filter (0.1–20 Hz) and a notch filter (60 Hz). Adaptive filtering (He et al. 2004) was used to correct for ocular artifacts, which were recorded from electrooculogram locations. ERPs were derived from Fz, Cz, and Pz channels and segmented (−100ms to 900 ms), baseline corrected, and averaged by stimulus condition. Brain vital sign ERP responses were manually verified by a blinded reviewer by identifying the relevant local maxima/minima within expected polarities and temporal ranges for the N100, the P300 and the N400. Peaks were then evaluated for both latency (response time) and amplitude (synchronous processing), for a total of six measures. The N400 amplitude was recorded as the mean voltage from 350-450 ms. N400 latency was recorded as the response time of the selected peak.

### King-Devick

K-D testing was completed in a quiet clinical setting both pre-season and post-season. Testing was conducted on an electronic tablet that was held at a comfortable reading distance. Players were instructed to read the numbers aloud as quickly and as accurately as possible from left to right and top to bottom. They completed an untimed demonstration card, followed by three timed test cards. The summed time of the three test cards was the player’s K-D score. At baseline, players completed the test twice and their best score was recorded. At post-season, the players completed the test once and their resultant K-D score was recorded.

### Blood Sampling

Non-fasting venous blood samples were collected from the antecubital fossa by standard venipuncture procedures. Samples were collected in Serum Separator Tubes containing a clot activator and serum gel separator. Serum samples were assayed on the same instrument and by the same person in a blinded fashion. NfL concentrations in plasma were measured with the NF-Light digital immunoassay (Quanterix, Cat#103186) using the HD-X Analyzer per the manufacturer’s protocol. In brief, samples were thawed on ice, mixed thoroughly by low-speed vortex and centrifuged at 10,000 g for 5 minutes before transferring samples to 96-well plates. Samples were diluted 1:4 by the instrument and tested in duplicate. In addition to participant serum samples, 8 calibrators and 2 quality control samples provided with the kits were included in the assay. Concentrations were interpolated from the standard curve using a 4-parameter logistic curve fit (1/y2 weighted).

### Supplementation

The CON group received a placebo while the SQ group received the supplement, Synaquell^TM^. The ingredients for Synaquell^TM^ and placebo supplements are listed in Supplemental Table 1. Athletes mixed one scoop (7.7 grams) of the provided powder with water and ingested twice daily. Supplements were administered by research team staff at team practices and home games. Supplements were prepared for players on road trips or days without organized team activities.

Research staff tracked all doses administered during practices and home games. Players self-reported all doses taken outside the supervision of the research staff using an internet-based or paper survey. All subjects and research staff responsible for data collection were blinded to the group assignment. Both groups discontinued the intervention after the final regular season hockey game. The NAHL played 60 games and NA3HL played 47 games during this study. Both teams participated in two 60-minute ice hockey games, and 4 or 5 practices lasting 60 to 90 minutes each week.

### Statistical Analysis

Statistical analyses were conducted using SPSS (IBM, New York, USA). Descriptive statistics were calculated for all dependent variables. T-tests were conducted to evaluate differences between groups across participant demographics for age, height, weight, number of games played, self-reported concussion history, and intervention compliance (as % of doses taken). The data from this study are available upon request from the corresponding author.

### Multivariate Analysis – Objective function and structure assessments

A repeated measures multivariate general linear analysis of variance (GLM) was used to investigate the effect of group allocation on dependent variables (6 brain vital sign measures, K-D, and NfL) as a factor of time (pre- to post- season). Prior concussion reporting and number of games played during the season were included as covariates. The number of prior concussions was derived from subjective participant recall as a standard aspect of the SCAT5 questionnaire, clinically diagnosed concussions during the season were not monitored as part of this study design. Post-hoc tests (both within and between subjects) were conducted to evaluate effects within individual dependent variables.

### Multivariate Analysis – Clinical Questionnaires

The statistical model design was then repeated using only the four clinical questionnaires (SCAT5, nQoL, PHQ-4, PGIC) as the dependent variables. PGIC scores for all participants at pre-season was set to 0, given that it is a retrospective analysis of change.

### Brain vital signs univariate analysis

To compare with prior evaluations of pre- to post-season changes of brain vital signs in contact athletes, grand average ERP waveforms (means +- 95% confidence intervals) were generated for deviant tone and incongruent word responses for each group. Paired, two-tailed Welch’s t-tests were performed on regions of interest identified on the waveforms as areas of deviation between the confidence intervals. Standardized radar plots were also generated for brain vital signs to indicate relative changes in scores over time.

## Results

### Participants

Of the 28 players randomized into the CON group, five players withdrew, seven left their team, and one was injured reducing the sample size to N=15. Of the 26 players randomized into the SQ group, three withdrew, six left their team, one was injured, and one dataset was compromised reducing the sample size to N=15. Due to supply chain delays, the intervention began 105 days after pre-season testing of the NA3HL team and 90 days after pre-season testing of the NAHL team. By this time, 26 NAHL games and 25 NA3HL games had occurred. Players were supplemented for the remaining 84 days (22 games) of the NA3HL season and 112 days (35 games) of the NAHL season. Postseason testing occurred 189 days after pre-season testing for the NA3HL team and 202 days for the NAHL team. There were no significant differences between groups across all demographics (Table 1).

**Table 1:** Player Demographics

| | Both Groups<br>(Mean ± SD) | CON<br>(Mean ± SD) | SQ<br>(Mean ± SD) | Between Group T-test<br>T-score (p value) |
| --- | --- | --- | --- | --- |
| Age | 18.73 ± 0.69 | 18.87 ± 0.74 | 18.60 ± 0.63 | 1.058 (0.299) |
| Height (cm) | 185.27 ± 7.24 | 187.27 ± 8.59 | 183.27 ± 5.13 | 1.552 (0.134) |
| Weight (kg) | 82.51 ± 9.26 | 84.93 ± 10.17 | 80.09 ± 7.86 | 1.458 (0.157) |
| Games Played | 39.03 ± 13.91 | 40.20 ± 12.94 | 37.87 ± 15.18 | 0.453 (0.654) |
| Concussion History | 1.00 ± 1.49 | 1.27 ± 1.91 | 0.73 ± 0.88 | 0.983 (0.338) |
| Compliance (% doses) | 84.42 ± 10.70 | 83.19 ± 11.77 | 85.64 ± 9.78 | -0.620 (0.541) |

### Multivariate Analysis – Objective assessments

The multivariate repeated-measures general linear model (Table 2) showed a significant overall within-subjects interaction effect of Group with Time (p = 0.036), which identified differential changes between the CON and SQ groups from pre- to post-season across all dependent variables. Post-hoc tests identified specific significant differences in both N100 latency (p = 0.005) and K-D score (p = 0.043).

**Table 2:** Repeated Measures General Linear Model Results–- Objective Tests

| Multivariate Tests |  |  |  |  |  |  |
| --- | --- | --- | --- | --- | --- | --- |
| Effect |  | Wil's Lambda | F | Hypothesis df | Error df | Sig |
| Within Subjects | Time | 0.503 | 2.347 | 8 | 19 | 0.061 |
|  | Time * Concussion Hx | 0.304 | 5.426 | 8 | 19 | **0.001\*\*** |
|  | Time * Games played | 0.501 | 2.366 | 8 | 19 | 0.059 |
|  | Time * Group | 0.467 | 2.706 | 8 | 19 | **0.036\*** |
| Between subjects | Intercept | 0.022 | 104.576 | 8 | 19 | **<.001\*\*\*** |
|  | Concussion history | 0.512 | 2.263 | 8 | 19 | 0.069 |
|  | Games played | 0.868 | 0.361 | 8 | 19 | 0.928 |
|  | Group | 0.729 | 0.882 | 8 | 19 | 0.549 |

Post-Hoc Tests of Within-Subject Effects
|  | Time |  | Time * Concussion history |  | Time * Games played |  | Time * Group |  |
| --- | --- | --- | --- | --- | --- | --- | --- | --- |
|  | F | Sig | F | Sig | F | Sig | F | Sig |
| N100 amplitude | 0.015 | 0.904 | 0.868 | 0.360 | 0.020 | 0.887 | 0.289 | 0.595 |
| N100 latency | 0.045 | 0.833 | 4.394 | **0.046\*** | 0.869 | 0.360 | 9.261 | **0.005\*\*** |
| P300 amplitude | 0.000 | 0.999 | 1.197 | 0.284 | 0.512 | 0.481 | 0.396 | 0.535 |
| P300 latency | 0.221 | 0.642 | 3.161 | 0.087 | 0.705 | 0.409 | 1.205 | 0.282 |
| N400 amplitude | 3.032 | 0.093 | 0.312 | 0.581 | 1.120 | 0.300 | 2.993 | 0.095 |
| N400 latency | 2.858 | 0.103 | 8.562 | **0.007\*\*** | 5.875 | **0.023\*** | 2.884 | 0.101 |
| King-Devick | 12.310 | **0.002\*\*** | 3.122 | 0.089 | 2.486 | 0.127 | 4.523 | **0.043\*** |
| NfLScore | 0.963 | 0.336 | 7.204 | **0.012\*** | 1.171 | 0.289 | 3.899 | 0.059 |

Post-Hoc Tests of Between-Subject Effects
|  | Intercept |  | Concussion history |  | Games played |  | Group |  |
| --- | --- | --- | --- | --- | --- | --- | --- | --- |
|  | F | Sig | F | Sig | F | Sig | F | Sig |
| N100 amplitude | 17.382 | **<.001\*\*\*** | 0.462 | 0.503 | 0.008 | 0.929 | 0.029 | 0.867 |
| N100 latency | 188.844 | **<.001\*\*\*** | 0.374 | 0.546 | 0.260 | 0.614 | 1.571 | 0.221 |
| P300 amplitude | 37.080 | **<.001\*\*\*** | 6.230 | **0.019\*** | 0.234 | 0.632 | 1.906 | 0.179 |
| P300 latency | 250.560 | **<.001\*\*\*** | 5.333 | **0.029\*** | 0.069 | 0.795 | 2.847 | 0.103 |
| N400 amplitude | 19.047 | **<.001\*\*\*** | 1.649 | 0.210 | 0.417 | 0.524 | 0.002 | 0.961 |
| N400 latency | 377.801 | **<.001\*\*\*** | 5.383 | **0.028\*** | 1.182 | 0.287 | 1.464 | 0.237 |
| King-Devick | 78.686 | **<.001\*\*\*** | 0.122 | 0.729 | 1.443 | 0.241 | 0.521 | 0.477 |
| NfL Score | 37.279 | **<.001\*\*\*** | 2.171 | 0.153 | 0.011 | 0.919 | 0.126 | 0.725 |

Group is modeled as the between-subjects factor and time point is the within-subjects factor. Games played and concussion history are included in the model as covariates.
\* $p < 0.05$ , \*\* $p < 0.01$ , and \*\*\* $p < 0.001$ . Bold values are where $p < 0.05$ .

There was also a significant, multivariate within-subjects effect between Time and Concussion History (p = 0.001). Post-hoc tests showed that the number of self-reported concussions, independent of group allocation, was significantly related to changes in outcome measures over time for the N100 latency (p = 0.046), N400 latency (p = 0.007), and NfL score (p = 0.012). There were no significant multivariate effects between subjects.

### Multivariate Analysis – Clinical Questionnaires

The multivariate repeated-measures general linear model for the clinical questionnaires (Table 3) showed no significant effects of group by time (p = 0.892). There were no significant effects or interaction effects for the covariates at any level of the model.

**Table 3:** Repeated Measures General Linear Model Results – Clinical Questionnaires

| Multivariate Tests |  |  |  |  |  |  |
| --- | --- | --- | --- | --- | --- | --- |
| Effect |  | Wil's Lambda | F | Hypothesis df | Error df | Sig |
| Within Subjects | Time | .675 | 2.522 | 4 | 21 | .072 |
|  | Time * Concussion Hx | .939 | .344 | 4 | 21 | .845 |
|  | Time * Games played | .858 | .870 | 4 | 21 | .498 |
|  | Time * Group | .950 | .274 | 4 | 21 | .892 |
| Between subjects | Intercept | .143 | 31.342 | 4 | 21 | **<.001\*\*** |
|  | Concussion history | .779 | 1.487 | 4 | 21 | .242 |
|  | Games Played | .930 | .393<sup>b</sup> | 4 | 21 | .811 |
|  | Group | .825 | 1.115 | 4 | 21 | .376 |

| Post-Hoc Tests of Within-Subject Effects |  |  |  |  |  |  |  |  |
| --- | --- | --- | --- | --- | --- | --- | --- | --- |
|  | Time |  | Time * Concussion history |  | Time * Games played |  | Time * Group |  |
|  | F | Sig | F | Sig | F | Sig | F | Sig |
| PGIC | 5.989 | **.022** | .125 | .727 | .994 | .329 | .125 | .726 |
| SCAT 5 | 1.867 | .184 | .200 | .659 | 1.810 | .191 | .313 | .581 |
| PHQ 4 | .158 | .694 | .167 | .686 | .214 | .648 | .262 | .613 |
| nQoL | 1.220 | .280 | 1.242 | .276 | .155 | .698 | .464 | .502 |

| Post-Hoc Tests of Between-Subject Effects |  |  |  |  |  |  |  |  |
| --- | --- | --- | --- | --- | --- | --- | --- | --- |
|  | Intercept |  | Concussion history |  | Games played |  | Group |  |
|  | F | Sig | F | Sig | F | Sig | F | Sig |
| PGIC | 5.989 | **.022\*** | .125 | .727 | .994 | .329 | .125 | .726 |
| SCAT 5 | 2.052 | .165 | .276 | .604 | .495 | .489 | .022 | .882 |
| PHQ 4 | 1.389 | .250 | 2.233 | .148 | .330 | .571 | 1.080 | .309 |
| nQoL | 56.068 | **<.001\*\*** | 2.771 | .109 | .112 | .741 | .000 | .987 |

Group is modeled as the between-subjects factor and time point is the within-subjects factor. Games played and concussion history are included in the model as covariates.
\* $p < 0.05$ , \*\* $p < 0.01$ , and \*\*\* $p < 0.001$ . Bold values are where $p < 0.05$ .

**Table 4:** Univariate t-tests from pre-season to post-season on brain vital signs waveform data

|  |  | t | df | 95% Confidence Interval |  |  |
| --- | --- | --- | --- | --- | --- | --- |
|  |  |  |  | sig | Low | High |
| N100 | CON | -2.156 | 14 | **0.049\*** | -21.81 | .0584 |
| Latency | SQ | 1.576 | 14 | 0.137 | -2.551 | 16.685 |
| N400 | CON | 2.715 | 14 | **0.017\*** | 0.184 | 1.566 |
| Amplitude | SQ | 0.682 | 14 | 0.506 | -0.340 | 0.658 |

Violin plots compare the objective measures and clinical questionnaires as a function of pre- versus post-difference for the CON and the SQ groups (Figure 1). The plot enables examination of the distributions of within-subject changes, separated by test type. Best-fit CON and SQ group differences are also line plotted with associated p-values for time effects from the respective multivariate GLM post-hoc tests. The violin plots show distribution differences between the CON and SQ groups in terms of pre- versus post-changes and highlight sensitivity differences between objective and subjective tests.

**Figure 1:**
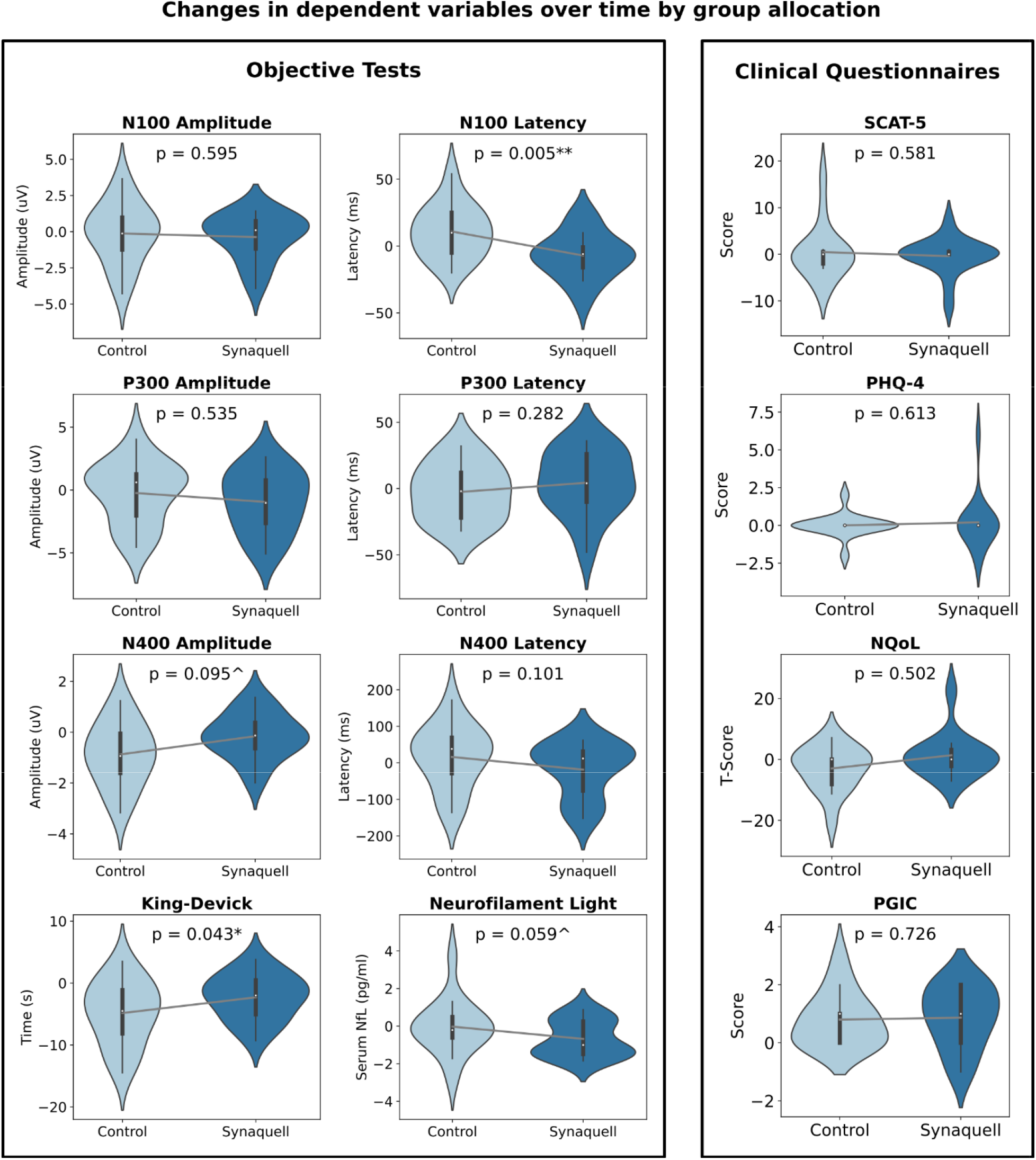
Violin plots showing pre- versus post- season differences in CON (light blue) and SQ (dark blue) groups. Plots are separated by test type showing objective measures (left side) and subjective measures (right side). Lines connecting plots show best fit differences with statistical difference above (* P < 0.05, ** P < 0.01). Note the statistical differences are from post-hoc tests in the multivariate GLM, which do not show significant effects from other tests (e.g., N400 amplitude reduction in brain vital sign waveform analysis). Examination of the violin plots show distribution differences between CON and SQ, along with sensitivity differences between objective and subjective tests. SCAT-5: The Sport Concussion Assessment Tool 5, PHQ-4: Patient Health Questionnaire-4, nQoL: The Neuro-Quality of Life Short Form Version 2.0 – Cognitive Function, and PGIC: Patient Global Impression of Change questionnaire.

### Brain vital signs waveform analysis

Grand average NeuroCatch ERP waveforms (+- 95CI of the mean) for CON (left) and SQ (right) groups were created for both deviant tone and congruent word stimuli (Figure 2). Regions of interest were identified for both the N100 latency, and the N400 amplitude components. Subsequent t-tests at each group identified significant delay in the N100 latency (p = 0.048) in the CON group, but not the SQ group (p = 0.137). Similarly, there were significant reductions in N400 amplitude (p = 0.017) in the CON group, which was not present in the SQ group (p = 0.506). Standardized brain vital sign radar plots compare CON (left) and SQ (right) with pre- versus post-season comparisons for both groups (Figure 3).

**Figure 2:**
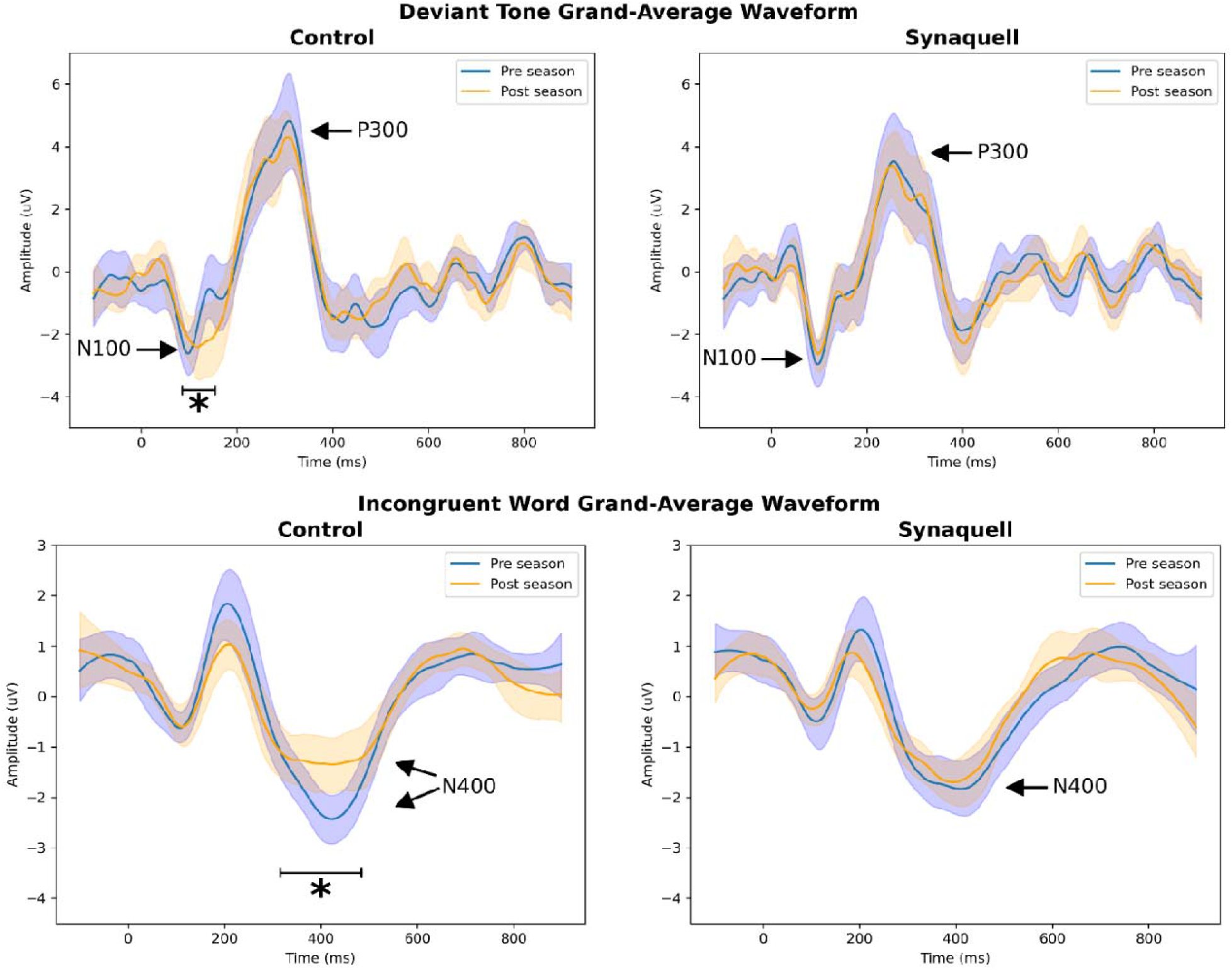
Grand average NeuroCatch ERP waveforms (+- 95CI of the mean) for CON (left) and SQ (right) groups (*p <0.05). Pre-season is in blue and post-season is in orange. The N100 and P300 to deviant tone stimuli (top row) and the N400 to incongruent word stimuli (bottom row). Time is on the x-axis (ms) and voltage is on the y-axis (µVs). Note the N100 latency delay and the N400 amplitude reduction in the CON group that is not present in the SQ group.

**Figure 3:**
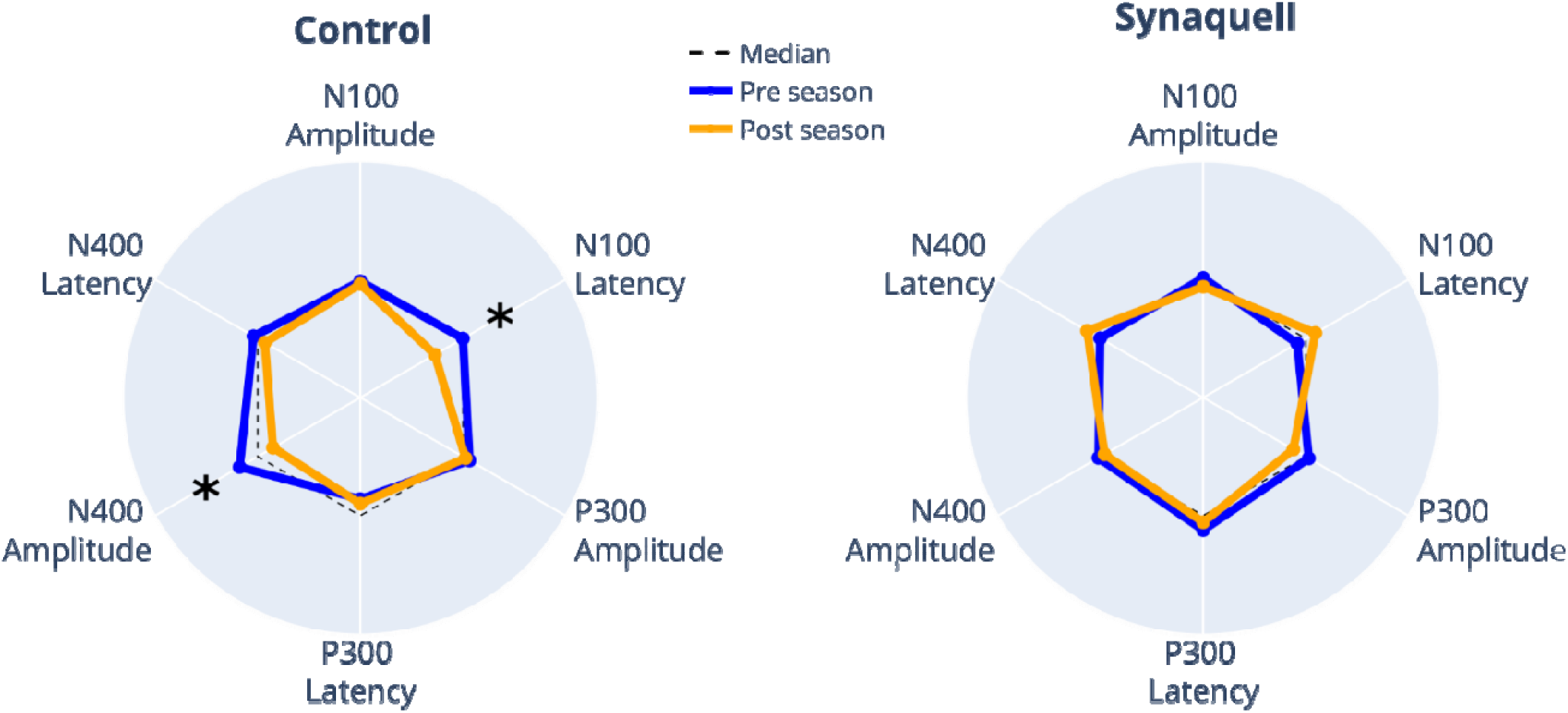
Standardized NeuroCatch^®^ radar plots showing brain vital sign results for each group at each time point (*p < 0.05). All 6 brain vital sign measures are plotted for both the CON (left) and SQ (right) groups. Pre-season results are in blue and post-season results are in orange. The dashed line denotes the median from prior normative data. Note the CON results replicate prior subconcussion impairment findings (Fickling et al., 2019, 2021a,b).

## Discussion

### Main findings

The aim of this study was to investigate the effects of the dietary supplement Synaquell^TM^ on brain function and structure. Multivariate analysis, including brain vital signs, K-D, NfL, showed significant within-subject differences between groups in changes from pre-to post-season (p = 0.036) (Table 2, Figure 1; Primary Hypothesis). Post-hoc tests revealed significant differential changes between groups in functional variables, including N100 latency and King-Devick scores. Specifically, the N100 latency became slower in the CON group and faster in the SQ group from pre-season to post-season (Figure 1, Supplemental Table 2). While K-D scores in both groups improved from pre-season to post-season, the improvement in the CON group was faster (Figure 1, Supplemental Table 2).

Additionally, a significant within-subject multivariate effect (p = 0.001) was observed for Concussion History * Time. Post-hoc testing identified significant changes in functional measures of N100 latency, N400 latency, and the structural NfL measure. This suggests that increased head impact exposure is linked to greater changes in both brain function and structure over the course of the season, regardless of group allocation.

The secondary analysis replicated previous findings on neurocognitive changes in athletes with no intervention (i.e., the CON group), specifically the delay in the N100 latency and decrease in N400 amplitude (Figure 3, Secondary Hypothesis). In contrast, no significant change was observed in the SQ group from pre- to post-season (Figs. 2&3).

Differential changes in objective brain function measures between the CON and SQ subjects from pre- to post-season supports the use of the dietary supplement Synaquell^TM^, to support brain function and structure in healthy, male athletes. To our knowledge, this is the first human study in a population of healthy, non-concussed athletes in which a multi-ingredient dietary supplement was found to be effective in supporting neurocognitive function and brain structure (Feinberg et al. 2023). The most similarly advanced studies in such a population have investigated Omega-3 Fatty Acids in relation to brain structure (Oliver et al. 2016; Heileson et al 2021; Mullins et al. 2022). Our results coincide with the findings of Oliver et al. and Heileson et al., in which serum NfL was attenuated by the daily administration of DHA in athletes (Oliver et al. 2016; Heileson et al. 2021). Mullins et al. 2022 alternatively found that daily DHA supplementation did not reduce serum NfL levels over the course of the season (Mullins et al. 2022). Studies examining the effects of single-ingredient dietary supplementation on neurocognition and brain structure have been conducted in concussed populations post-injury (Feinberg et al. 2023). Multiple ingredients have the potential to impact a variety of pathways and processes in the neurometabolic cascade, which may improve effectiveness (Lucke-Wold et al. 2016).

### Brain vital signs

The current study provides the latest evidence that head impact exposure affects cognitive processing (N400) and auditory sensory processing (N100). A pattern involving the N100 and N400 is emerging across different ages and sports. Specific responses vary slightly in terms of amplitude, latency and waveform shape across studies, but the consistency of N100 and N400 impairment in athletes exposed to repetitive head impacts is notable (Fickling et al. 2019, 2021a,b).

While the P300 component has shown little change to-date when examining group average data, the results of the multivariate analyses indicate greater complexity with respect to individual variability and factors affecting head impact exposure. For instance, the P300 amplitude and latency, as markers of basic attention, co-varied significantly with subjective concussion history (as did N400 latency). Similarly, significant covariance occurred between the N100 and the K-D test for pre- versus post-comparisons by group by games played (as did the N400 for pre- versus post by games played). Of interest, NfL results significantly co-varied in terms of pre- versus post-season and subjective concussion history. Taken together, the current findings suggest that while N100 and N400 changes may serve as an initial indicator of subconcussive impairment, additional brain vital sign features may also be informative as objective markers for underlying sub-concussive impairment profiles across a spectrum of differential head-impact exposure between players.

### Objective assessments of function and structure

There were no significant differences between the groups in clinical neuropsychological questionnaires pre- to post-season despite significant differences in objective neurocognitive measures. This observation is not surprising, given that clinical questionnaires such as the SCAT5 are subjective in nature, rely heavily upon symptom reporting, and healthy, non-concussed athletes that are exposed to repetitive head impacts typically do not show observable physical signs of neurological impairment (Lember et al. 2021). The SCAT5 has been shown to differentiate concussed and non-concussed athletes in an acute concussion diagnosis setting; however, it is less useful in monitoring recovery (Echmendia et al. 2017). The other neuropsychological questionnaires (PHQ-4, NQoL, and the PGIC) also did not show differences between groups that were detected by objective measures. Nonetheless, these questionnaires provide a subjective, yet insightful glimpse into the functional state of the individual’s brain health.

The current findings highlight the importance of expanding upon studies investigating the relationship between dietary supplementation, cognitive function and brain structure in athletes exposed to repetitive head impacts. Also important to note is the relationship between objective measures of concussion and subjective clinical symptoms. Batteries of objective tools such as the brain vital signs, K-D, and NfL may be able to detect neurocognitive impairments that are at or below current diagnostic thresholds for concussion. A critical issue involves the point at which cognitive impairments manifest as detectable clinical-behavioral symptoms. Further investigation is needed to investigate if dietary supplements alter or prevent the emergence of clinical-behavioral symptomatology.

### Limitations

This interventional trial has several limitations. First, the cohort was exclusively male Junior-A hockey players; therefore, further studies are required to generalize the results. These investigations should also examine factors related to concussion diagnosis, including gender, age, and mechanism of injury (e.g., blast injuries). Second, variables related to the specific dietary supplement, Synaquell^TM^, need further analysis. Synaquell^TM^ doses were not standardized by body weight, and individuals with higher body mass may require higher dosage to experience the same effects on neurocognition. Future studies should evaluate optimal dosages, ingredients, and the potential for individually tailored dietary supplementation. Third, supplementation did not begin at the beginning of the season due to supply chain challenges. The athletes participated in a portion of the season without receiving the supplement. Season long supplementation may have contributed to greater neurocognitive changes (or lack thereof) in the SQ group. Fourth, our study examined only pre- versus post- timelines, and further time points should be investigated in future studies, including whether dietary supplements support the brain during recovery from a clinically diagnosed concussion. Finally, the current study did not include quantified impact evaluation, so the relationship with impact exposure remains to be determined.

## Conclusion

This prospective, randomized trial showed that a multi-ingredient dietary supplement significantly affected objective measures of brain function and structure from pre- to post-season in Junior A ice hockey players compared to a placebo control group. Further investigation into the effects of dietary supplementation on the contact athlete’s brain is warranted.

## Data Availability

The data from this study are available upon request from the corresponding author.

## Acknowledgments

The current study is part of the clinical trial listed on ClinicalTrials.gov (ClinicalTrials.gov Identifier: NCT05498818). The study was funded by the USA Hockey Foundation (M. Stuart). The authors would like to thank Dr. Aynsley Smith for laying the foundation that made this research possible. We would like to thank Sydney Kalina, Chad Eickhoff, Jeff Lamb and Houston Hawkins for continued support with the implementation of data collection throughout this study. The authors would also like to thank Thorne for providing Synaquell^TM^ along with on-going implementation support throughout the clinical trial. The NeuroCatch^®^ Platform was provided by HealthTech Connex.

---

**Supplemental Table 1:** SynaquellTM Ingredients

| **Ingredient** | **Amount** |
| --- | --- |
| Beta Hydroxybutyrate (as Magnesium Beta Hydroxybutyrate) | 1000 mg |
| Glutathione (Reduced) | 500 mg |
| Nicotinamide Riboside Hydrogen Malate | 415 mg |
| Riboflavin (as Riboflavin-5-phosphate) | 50 mg |
| Magnesium (as Magnesium Beta Hydroxybutyrate) | 118 mg |
| Leucine | 2500 mg |
| Isoleucine | 1250 mg |
| Valine | 1250 mg |
| Resveratrol | 250 mg |
| MERIVA (Curcumin Phytosome) | 250 mg |
| Docosahexaenoic Acid (DHA) Vegan | 250 mg |
| Ubiquinone (Coenzyme Q10) | 100 mg |
| Betaine Anhydrous (Trimethylglycine) | 85 mg |

**Supplemental Table 2:** Descriptive Statistics (Mean ± SD) for dependent variables

|  | CON |  | SQ |  |
| --- | --- | --- | --- | --- |
|  | Pre-season | Post-Season | Pre-season | Post-Season |
| N100 amplitude | 3.28 ± 1.49 | 3.16 ± 1.87 | 3.42 ± 1.66 | 3.05 ± 1.27 |
| N100 latency | 98 ± 13.63 | 108.93 ± 21.37 | 101.33 ± 10.76 | 94.27 ± 14.66 |
| P300 amplitude | 5.32 ± 2.39 | 5.08 ± 1.94 | 5.1 ± 2.62 | 4.16 ± 2.1 |
| P300 latency | 260.27 ± 36 | 257.73 ± 30.83 | 242.93 ± 37.79 | 247.33 ± 32.09 |
| N400 amplitude | 2.19 ± 0.88 | 1.31 ± 1.00 | 1.77 ± 0.98 | 1.61 ± 0.82 |
| N400 latency | 408.13 ± 35.29 | 424.27 ± 72.43 | 413.47 ± 62.87 | 394.93 ± 46.43 |
| King-Devick | 47.34 ± 12.09 | 42.5 ± 10.35 | 43.5 ± 10.1 | 41.2 ± 8.11 |
| NfL | 4.77 ± 1.39 | 4.74 ± 1.43 | 5.13 ± 1.67 | 4.45 ± 1.33 |
| SCAT 5 | 3.47 ± 5.13 | 3.93 ± 7.71 | 3.13 ± 7.74 | 2.73 ± 5.06 |
| PHQ 4 | 0.67 ± 1.45 | 0.67 ± 1.45 | 1.07 ± 1.91 | 1.27 ± 2.60 |
| NQoL | 23.81 ± 5.63 | 20.83 ± 8.02 | 21.41 ± 8.34 | 21.99 ± 5.86 |
| PGIC | / | 0.80 ± 0.94 | / | 0.87 ± 1.06 |

## References

Bailes JE, Petraglia AL, Omalu BI, Nauman E, Talavage T. Role of subconcussion in repetitive mild traumatic brain injury. J Neurosurg. 2013 Nov;119(5):1235–45. doi: 10.3171/2013.7.JNS121822. Epub 2013 Aug 23. PMID: 23971952.

Mainwaring L, Ferdinand Pennock KM, Mylabathula S, Alavie BZ. Subconcussive head impacts in sport: A systematic review of the evidence. Int J Psychophysiol. 2018 Oct;132(Pt A):39–54. doi: 10.1016/j.ijpsycho.2018.01.007. Epub 2018 Feb 3. PMID: 29402530.

Abbas K, Shenk TE, Poole VN, Breedlove EL, Leverenz LJ, Nauman EA, Talavage TM, Robinson ME. Alteration of default mode network in high school football athletes due to repetitive subconcussive mild traumatic brain injury: a resting-state functional magnetic resonance imaging study. Brain Connect. 2015 Mar;5(2):91–101. doi: 10.1089/brain.2014.0279. Epub 2014 Oct 21. PMID: 25242171.

Talavage TM, Nauman EA, Breedlove EL, Yoruk U, Dye AE, Morigaki KE, Feuer H, Leverenz LJ. Functionally-detected cognitive impairment in high school football players without clinically-diagnosed concussion. J Neurotrauma. 2014 Feb 15;31(4):327–38. doi: 10.1089/neu.2010.1512. Epub 2013 Apr 11. PMID: 20883154; PMCID: PMC3922228.

Tsushima WT, Geling O, Arnold M, Oshiro R. Are There Subconcussive Neuropsychological Effects in Youth Sports? An Exploratory Study of High- and Low-Contact Sports. Appl Neuropsychol Child. 2016;5(2):149–55. doi: 10.1080/21622965.2015.1052813. Epub 2016 Mar 15. Erratum in: Appl Neuropsychol Child. 2016 Oct-Dec;5(4):311. PMID: 26979930.

Mez J, Daneshvar DH, Abdolmohammadi B, Chua AS, Alosco ML, Kiernan PT, Evers L, Marshall L, Martin BM, Palmisano JN, Nowinski CJ, Mahar I, Cherry JD, Alvarez VE, Dwyer B, Huber BR, Stein TD, Goldstein LE, Katz DI, Cantu RC, Au R, Kowall NW, Stern RA, McClean MD, Weuve J, Tripodis Y, McKee AC. Duration of American Football Play and Chronic Traumatic Encephalopathy. Ann Neurol. 2020 Jan;87(1):116–131. doi: 10.1002/ana.25611. Epub 2019 Nov 23. PMID: 31589352; PMCID: PMC6973077.

Pender, Sara C. MBA1; Smith, Aynsley M. PhD, RN2,3; Finnoff, Jonathan T. DO3; Huston, John III MD4; Stuart, Michael J. MD5. Concussions in Ice Hockey — Moving Toward Objective Diagnoses and Point-of-care Treatment: A Review. Current Sports Medicine Reports: September 2020 - Volume 19 - Issue 9 - p 380–386 doi: 10.1249/JSR.0000000000000752

Ghosh Hajra S, Liu CC, Song X, Fickling S, Liu LE, Pawlowski G, Jorgensen JK, Smith AM, Schnaider-Beeri M, Van Den Broek R, Rizzotti R, Fisher K, D’Arcy RC. Developing Brain Vital Signs: Initial Framework for Monitoring Brain Function Changes Over Time. Front Neurosci. 2016 May 12;10:211. doi: 10.3389/fnins.2016.00211. PMID: 27242415; PMCID: PMC4867677.

Davis, P. A. (1939). Effects of acoustic stimuli on the waking human brain. Journal of neurophysiology, 2(6), 494–499.

Sutton S, Tueting P, Zubin J, John ER. Information delivery and the sensory evoked potential. Science. 1967 Mar 17;155(3768):1436–9. doi: 10.1126/science.155.3768.1436. PMID: 6018511.

Kutas M, Hillyard SA. Reading senseless sentences: brain potentials reflect semantic incongruity. Science. 1980 Jan 11;207(4427):203–5. doi: 10.1126/science.7350657. PMID: 7350657.

Fickling SD, Smith AM, Stuart MJ, Dodick DW, Farrell K, Pender SC, D’Arcy RCN. Subconcussive brain vital signs changes predict head-impact exposure in ice hockey players. Brain Commun. 2021 Apr 6;3(2):fcab019. doi: 10.1093/braincomms/fcab019. PMID: 33855296; PMCID: PMC8023684.

Fickling SD, Poel DN, Dorman JC, D’Arcy RCN, Munce TA. Subconcussive changes in youth football players: objective evidence using brain vital signs and instrumented accelerometers. Brain Commun. 2021 Dec 15;4(2):fcab286. doi: 10.1093/braincomms/fcab286. PMID: 35291689; PMCID: PMC8914875.

Fickling SD, Smith AM, Pawlowski G, Ghosh Hajra S, Liu CC, Farrell K, Jorgensen J, Song X, Stuart MJ, D’Arcy RCN. Brain vital signs detect concussion-related neurophysiological impairments in ice hockey. Brain. 2019 Feb 1;142(2):255–262. doi: 10.1093/brain/awy317. PMID: 30649205; PMCID: PMC6351777.

Nowak MK, Bevilacqua ZW, Ejima K, et al. Neuro-Ophthalmologic Response to Repetitive Subconcussive Head Impacts: A Randomized Clinical Trial. JAMA Ophthalmol. 2020;138(4):350–357. doi:10.1001/jamaophthalmol.2019.6128

Munce TA, Dorman JC, Odney TO, Thompson PA, Valentine VD, Bergeron MF. Effects of youth football on selected clinical measures of neurologic function: a pilot study. J Child Neurol. 2014 Dec;29(12):1601–7. doi: 10.1177/0883073813509887. Epub 2013 Nov 21. PMID: 24272520.

Joseph JR, Swallow JS, Willsey K, Lapointe AP, Khalatbari S, Korley FK, Oppenlander ME, Park P, Szerlip NJ, Broglio SP. Elevated markers of brain injury as a result of clinically asymptomatic high-acceleration head impacts in high-school football athletes. J Neurosurg. 2018 Jul 3:1–7. doi: 10.3171/2017.12.JNS172386. Epub ahead of print. PMID: 29966462.

Krause DA, Hollman JH, Breuer LT, Stuart MJ. Validity Indices of the King-Devick Concussion Test in Hockey Players. Clin J Sport Med. 2022 May 1;32(3):e313–e315. doi: 10.1097/JSM.0000000000000938. Epub 2021 May 7. PMID: 34009786.

Eddy R, Goetschius J, Hertel J, Resch J. Test-Retest Reliability and the Effects of Exercise on the King-Devick Test. Clin J Sport Med. 2020 May;30(3):239–244. doi: 10.1097/JSM.0000000000000586. PMID: 32341291.

Caccese JB, Best C, Lamond LC, Difabio M, Kaminski TW, Watson D, Getchell N, Buckley TA. Effects of Repetitive Head Impacts on a Concussion Assessment Battery. Med Sci Sports Exerc. 2019 Jul;51(7):1355–1361. doi: 10.1249/MSS.0000000000001905. PMID: 30649104.

Papa L, Walter AE, Wilkes JR, Clonts HS, Johnson B, Slobounov SM. Effect of Player Position on Serum Biomarkers during Participation in a Season of Collegiate Football. J Neurotrauma. 2022 Oct;39(19-20):1339–1348. doi: 10.1089/neu.2022.0083. Epub 2022 Sep 1. PMID: 35615873; PMCID: PMC9529311.

Oliver JM, Jones MT, Kirk KM, Gable DA, Repshas JT, Johnson TA, Andréasson U, Norgren N, Blennow K, Zetterberg H. Serum Neurofilament Light in American Football Athletes over the Course of a Season. J Neurotrauma. 2016 Oct 1;33(19):1784–1789. doi: 10.1089/neu.2015.4295. Epub 2016 Mar 16. PMID: 26700106.

Oliver JM, Anzalone AJ, Stone JD, Turner SM, Blueitt D, Garrison JC, Askow AT, Luedke JA, Jagim AR. Fluctuations in blood biomarkers of head trauma in NCAA football athletes over the course of a season. J Neurosurg. 2018 May 29:1–8. doi: 10.3171/2017.12.JNS172035. Epub ahead of print. PMID: 29807487.

Wirsching A, Chen Z, Bevilacqua ZW, Huibregtse ME, Kawata K. Association of Acute Increase in Plasma Neurofilament Light with Repetitive Subconcussive Head Impacts: A Pilot Randomized Control Trial. J Neurotrauma. 2019 Feb 15;36(4):548–553. doi: 10.1089/neu.2018.5836. Epub 2018 Sep 4. PMID: 30019617.

Rubin LH, Tierney R, Kawata K, Wesley L, Lee JH, Blennow K, Zetterberg H, Langford D. NfL blood levels are moderated by subconcussive impacts in a cohort of college football players. Brain Inj. 2019;33(4):456–462. doi: 10.1080/02699052.2019.1565895. Epub 2019 Jan 11. PMID: 30776989.

Smith, Aynsley M. RN, PhD*; Stuart, Michael J. MD†; Roberts, William O. MD, MS‡; Dodick, David W. MD§; Finnoff, Jonathan T. DO¶; Jorgensen, Janelle K. BA; Krause, David A. PT, DSc**. Concussion in Ice Hockey: Current Gaps and Future Directions in an Objective Diagnosis. Clinical Journal of Sport Medicine: September 2017 - Volume 27 - Issue 5 - p 503–509 doi: 10.1097/JSM.0000000000000412

Nauman, Eric A., Thomas M. Talavage, and Paul S. Auerbach. “Mitigating the consequences of subconcussive head injuries.” Annual Review of Biomedical Engineering 22 (2020): 387–407.

Mishra S, Singh VJ, Chawla PA, Chawla V. Neuroprotective Role of Nutritional Supplementation in Athletes. Curr Mol Pharmacol. 2022;15(1):129–142. doi: 10.2174/1874467214666211209144721. PMID: 34886789.

Walrand S, Gaulmin R, Aubin R, Sapin V, Coste A, Abbot M. Nutritional factors in sport-related concussion. Neurochirurgie. 2021 May;67(3):255–258. doi: 10.1016/j.neuchi.2021.02.001. Epub 2021 Feb 11. PMID: 33582206.

Lucke-Wold BP, Logsdon AF, Nguyen L, Eltanahay A, Turner RC, Bonasso P, Knotts C, Moeck A, Maroon JC, Bailes JE, Rosen CL. Supplements, nutrition, and alternative therapies for the treatment of traumatic brain injury. Nutr Neurosci. 2018 Feb;21(2):79–91. doi: 10.1080/1028415X.2016.1236174. Epub 2016 Oct 5. PMID: 27705610; PMCID: PMC5491366.

Oliver JM, Anzalone AJ, Turner SM. Protection Before Impact: the Potential Neuroprotective Role of Nutritional Supplementation in Sports-Related Head Trauma. Sports Med. 2018 Mar;48(Suppl 1):39–52. doi: 10.1007/s40279-017-0847-3. PMID: 29368186; PMCID: PMC5790849.

McIntosh TK, Vink R, Yamakami I, Faden AI. Magnesium protects against neurological deficit after brain injury. Brain Res. 1989 Mar 20;482(2):252–60. doi: 10.1016/0006-8993(89)91188-8. PMID: 2784989.

Standiford L, O’Daniel M, Hysell M, Trigger C. A randomized cohort study of the efficacy of PO magnesium in the treatment of acute concussions in adolescents. Am J Emerg Med. 2021 Jun;44:419–422. doi: 10.1016/j.ajem.2020.05.010. Epub 2020 Jul 9. PMID: 33243533.

Lopez MS, Dempsey RJ, Vemuganti R. Resveratrol neuroprotection in stroke and traumatic CNS injury. Neurochem Int. 2015 Oct;89:75–82. doi: 10.1016/j.neuint.2015.08.009. Epub 2015 Aug 12. PMID: 26277384; PMCID: PMC4587342.

Lin CJ, Chen TH, Yang LY, Shih CM. Resveratrol protects astrocytes against traumatic brain injury through inhibiting apoptotic and autophagic cell death. Cell Death Dis. 2014 Mar 27;5(3):e1147. doi: 10.1038/cddis.2014.123. PMID: 24675465; PMCID: PMC3973229.

Cheng YH, Zhao JH, Zong WF, Wei XJ, Xu Z, Yuan Y, Jiang YF, Luo X, Wang W, Qu WS. Acute Treatment with Nicotinamide Riboside Chloride Reduces Hippocampal Damage and Preserves the Cognitive Function of Mice with Ischemic Injury. Neurochem Res. 2022 Aug;47(8):2244–2253. doi: 10.1007/s11064-022-03610-3. Epub 2022 May 19. PMID: 35585298

Wu A, Ying Z, Schubert D, Gomez-Pinilla F. Brain and spinal cord interaction: a dietary curcumin derivative counteracts locomotor and cognitive deficits after brain trauma. Neurorehabil Neural Repair. 2011 May;25(4):332–42. doi: 10.1177/1545968310397706. Epub 2011 Feb 22. PMID: 21343524; PMCID: PMC3258099.

Aoyama K. Glutathione in the Brain. Int J Mol Sci. 2021 May 9;22(9):5010. doi: 10.3390/ijms22095010. PMID: 34065042; PMCID: PMC8125908.

Raikes AC, Hernandez GD, Mullins VA, Wang Y, Lopez C, Killgore WDS, Chilton FH, Brinton RD. Effects of docosahexaenoic acid and eicosapentaoic acid supplementation on white matter integrity after repetitive sub-concussive head impacts during American football: Exploratory neuroimaging findings from a pilot RCT. Front Neurol. 2022 Sep 15;13:891531. doi: 10.3389/fneur.2022.891531. PMID: 36188406; PMCID: PMC9521411.

Oliver JM, Jones MT, Kirk KM, Gable DA, Repshas JT, Johnson TA, Andréasson U, Norgren N, Blennow K, Zetterberg H. Effect of Docosahexaenoic Acid on a Biomarker of Head Trauma in American Football. Med Sci Sports Exerc. 2016 Jun;48(6):974–82. doi: 10.1249/MSS.0000000000000875. PMID: 26765633.

Heileson JL, Anzalone AJ, Carbuhn AF, Askow AT, Stone JD, Turner SM, Hillyer LM, Ma DWL, Luedke JA, Jagim AR, Oliver JM. The effect of omega-3 fatty acids on a biomarker of head trauma in NCAA football athletes: a multi-site, non-randomized study. J Int Soc Sports Nutr. 2021 Sep 27;18(1):65. doi: 10.1186/s12970-021-00461-1. PMID: 34579748; PMCID: PMC8477477.

Lee DC, Vali K, Baldwin SR, Divino JN, Feliciano JL, Fequiere JR, Fernandez MA, Frageau JC, Longo FK, Madhoun SS, Mingione V P, O’Toole TR, Ruiz MG, Tanner GR. Dietary Supplementation With the Ketogenic Diet Metabolite Beta-Hydroxybutyrate Ameliorates Post-TBI Aggression in Young-Adult Male *Drosophila*. Front Neurosci. 2019 Oct 30;13:1140. doi: 10.3389/fnins.2019.01140. PMID: 31736687; PMCID: PMC6833482.

Pierce JD, Shen Q, Peltzer J, Thimmesch A, Hiebert JB. A pilot study exploring the effects of ubiquinol on brain genomics after traumatic brain injury. Nurs Outlook. 2017 Sep-Oct;65(5S):S44–S52. doi: 10.1016/j.outlook.2017.06.012. Epub 2017 Jul 1. PMID: 28755974.

Pierce J.D., Gupte R., Thimmesch A., Shen Q., Hiebert J.B., Brooks W.M., Clancy R.L., Diaz F.J., Harris J.L. Ubiquinol treatment for TBI in male rats: Effects on mitochondrial integrity, injury severity, and neurometabolism. J. Neurosci. Res. 2018;96:1080–1092. doi: 10.1002/jnr.24210.

Aquilani R, Iadarola P, Contardi A, Boselli M, Verri M, Pastoris O, Boschi F, Arcidiaco P, Viglio S. Branched-chain amino acids enhance the cognitive recovery of patients with severe traumatic brain injury. Arch Phys Med Rehabil. 2005 Sep;86(9):1729–35. doi: 10.1016/j.apmr.2005.03.022. PMID: 16181934.

Giza CC, Hovda DA. The new neurometabolic cascade of concussion. Neurosurgery. 2014 Oct;75 Suppl 4(0 4):S24–33. doi: 10.1227/NEU.0000000000000505. PMID: 25232881; PMCID: PMC4479139.

He, P., Wilson, G. & Russell, C. Removal of ocular artifacts from electro-encephalogram by adaptive filtering. Med. Biol. Eng. Comput. 42, 407–412 (2004). https://doi.org/10.1007/BF02344717

Mullins VA, Graham S, Cummings D, Wood A, Ovando V, Skulas-Ray AC, Polian D, Wang Y, Hernandez GD, Lopez CM, Raikes AC, Brinton RD, Chilton FH. Effects of Fish Oil on Biomarkers of Axonal Injury and Inflammation in American Football Players: A Placebo-Controlled Randomized Controlled Trial. Nutrients. 2022 May 20;14(10):2139. doi: 10.3390/nu14102139. PMID: 35631280; PMCID: PMC9146417.

King D, Hume P, Gissane C, Clark T. Use of the King-Devick test for sideline concussion screening in junior rugby league. J Neurol Sci. 2015 Oct 15;357(1-2):75–9. doi: 10.1016/j.jns.2015.06.069. Epub 2015 Jul 2. PMID: 26152829.

Lember LM, Ntikas M, Mondello S, Wilson L, Hunter A, Di Virgilio T, Santoro E, Ietswaart M. Effects of sport-related repetitive subconcussive head impacts on biofluid markers: a scoping review protocol. BMJ Open. 2021 Jun 28;11(6):e046452. doi: 10.1136/bmjopen-2020-046452. PMID: 34183343; PMCID: PMC8240577.

Echemendia, R. J., Meeuwisse, W., McCrory, P., Davis, G. A., Putukian, M., Leddy, J., … & Herring, S. (2017). The sport concussion assessment tool 5th edition (SCAT5): background and rationale. British journal of sports medicine, 51(11), 848–850.

Feinberg C, Mayes KD, Jarvis R, Carr C, Mannix R. Nutritional supplement and dietary interventions as a prophylaxis or treatment of sub-concussive repetitive head impact (SRHI) and mild traumatic brain injury (mTBI): A systematic review. J Neurotrauma. 2023 Jan 21. doi: 10.1089/neu.2022.0498. Epub ahead of print. PMID: 36680752.

